# Integration of DNA methylation datasets for individual prediction

**DOI:** 10.1101/2023.03.22.23287572

**Authors:** Charlotte Merzbacher, Barry Ryan, Thibaut Goldsborough, Robert F Hillary, Archie Campbell, Lee Murphy, Andrew M McIntosh, David Liewald, Sarah E Harris, Allan F McRae, Simon R Cox, Timothy I Cannings, Catalina Vallejos, Daniel L McCartney, Riccardo E Marioni

## Abstract

**Background:** Epigenetic scores (EpiScores) can provide blood-based biomarkers of lifestyle and disease risk. Projecting a new individual onto a reference panel would aid precision medicine and risk communication but is challenging due to the separation of technical and biological sources of variation with array data. Normalisation methods can standardize data distributions but may also remove population-level biological variation.

**Methods:** We compared two independent birth cohorts (Lothian Birth Cohorts of 1921 and 1936 – n_LBC1921_ = 387 and n_LBC1936_ = 498) with DNA methylation assessed at the same chronological age (79 years) and processed in the same lab but in different years and experimental batches. We examined the effect of 15 normalisation methods on a BMI EpiScore (trained in an external cohort of 18,413 individuals) when the cohorts were normalised separately and together.

**Results:** The BMI EpiScore explained a maximum variance of R^2^=24.5% in BMI in LBC1936 after SWAN normalisation. Although there were differences in the variance explained across cohorts, the normalisation methods made minimal differences to the estimates within cohorts. Conversely, a range of absolute differences were seen for individual-level EpiScore estimates when cohorts were normalised separately versus together. While within-array methods result in identical BMI EpiScores whether a cohort was normalised on its own or together with the second dataset, a range of differences were observed for between-array methods.

**Conclusions:** Using normalisation methods that give similar EpiScores whether cohorts are analysed separately or together will minimise technical variation when projecting new data onto a reference panel. These methods are especially important for cases where when raw data and joint normalisation of cohorts is not possible or is computationally expensive.

## Introduction

There is an increasing focus on the application of epigenetic biomarkers in large cohort studies for health research [1]. For example, DNA methylation (DNAm)-based predictors – epigenetic scores or EpiScores – of adiposity, smoking, alcohol consumption (traits which typically suffer from measurement error due to recall bias), and protein levels may help to stratify individuals into risk groups and predict disease outcomes [2,3].

However, DNAm data sourced from different populations and lab environments can have technical and biological variation that is difficult to partition. Normalising DNAm datasets can account for technical variation but may remove meaningful biological variation. Understanding the impact of normalisation is vital if future studies are to integrate multiple methylation datasets or project new individuals onto existing datasets. Recent work has also described heterogeneity when applying different normalisation methods to replicate samples from the same individual [4]. That study showed considerable variation in the normalisation pipeline that yielded the highest intraclass correlations for 41 different EpiScores.

The Illumina Infinium HumanMethylation450 and EPIC arrays assess methylation genome-wide [5,6] and are widely used by cohort studies. Following a whole-genome amplification step, probes hybridize to target CpG sites and fluorescent markers signal methylation status. The arrays measure methylation using two probe types (Type I and Type II). Type I probes have two 50bp probes for each CpG site, one of which hybridizes to the methylated site (M) and the other to the unmethylated site (U). Type II probes are a single probe with two different dye colours to differentiate between M and U states.

Quantile normalisation (QN) is a nonlinear transformation that ensures the array-wide distributions of CpG values are identical by replacing the raw CpG values with the mean of all CpG features with the same rank [6]. QN can be used to correct bias due to differences between methylated and unmethylated dye intensities (dye bias correction) and bias due to Type I and Type II probe differences (between-array normalisation). In addition, background adjustment can control for the offset between Type I and Type II probe intensities. In addition to QN, which utilises information across samples, within-array (i.e. sample-indepedent) approaches also exist. These approaches include subset within-array normalisation (SWAN), which reduces the difference in distributions between Type I and Type II probes, based on a random subset of biologically similar probes [7]; beta-mixture quantile (BMIQ) normalisation, which performs adjustment on Type II probes, transforming their distribution to one more similar to Type I probes [8]; peak-based correction (PBC), a correction method which rescales Type II probe distributions on the basis of Type I probe data [9]; and normal-exponential out-of-band (Noob) normalisation, which performs background correction and dye-bias equalisation [10].

If technical noise across different datasets can be accounted for, new samples or datasets can be normalised individually rather than re-normalising all data together, which is computationally expensive. Here, we apply various quantile normalisation methods to two independent cohorts of age-matched older adults (considered separately and jointly) to determine which approach performs best for the projection of an individual or individuals onto a reference dataset.

## Results

Fifteen normalisation approaches [7,10,11] were ranked among three datasets (the Lothian Birth Cohort of 1921 (LBC1921), the Lothian Birth Cohort of 1936 (LBC1936), and LBC1921 + LBC1936 combined; **Figure 1**) [12]. Methylation was assessed on the Illumina 450k array in two separate experiments. First, samples from 387 individuals from LBC1921 taken between 1999 and 2001 at mean age 79.1 (SD = 0.58) years were processed. Second, samples from 498 individuals from LBC1936 taken between 2014 and 2017 at mean age 79.3 (SD = 0.62) years were processed. The combined cohort contained 885 individuals. Pre-normalisation filtering steps are described in the **Methods**.

**Figure 1:**
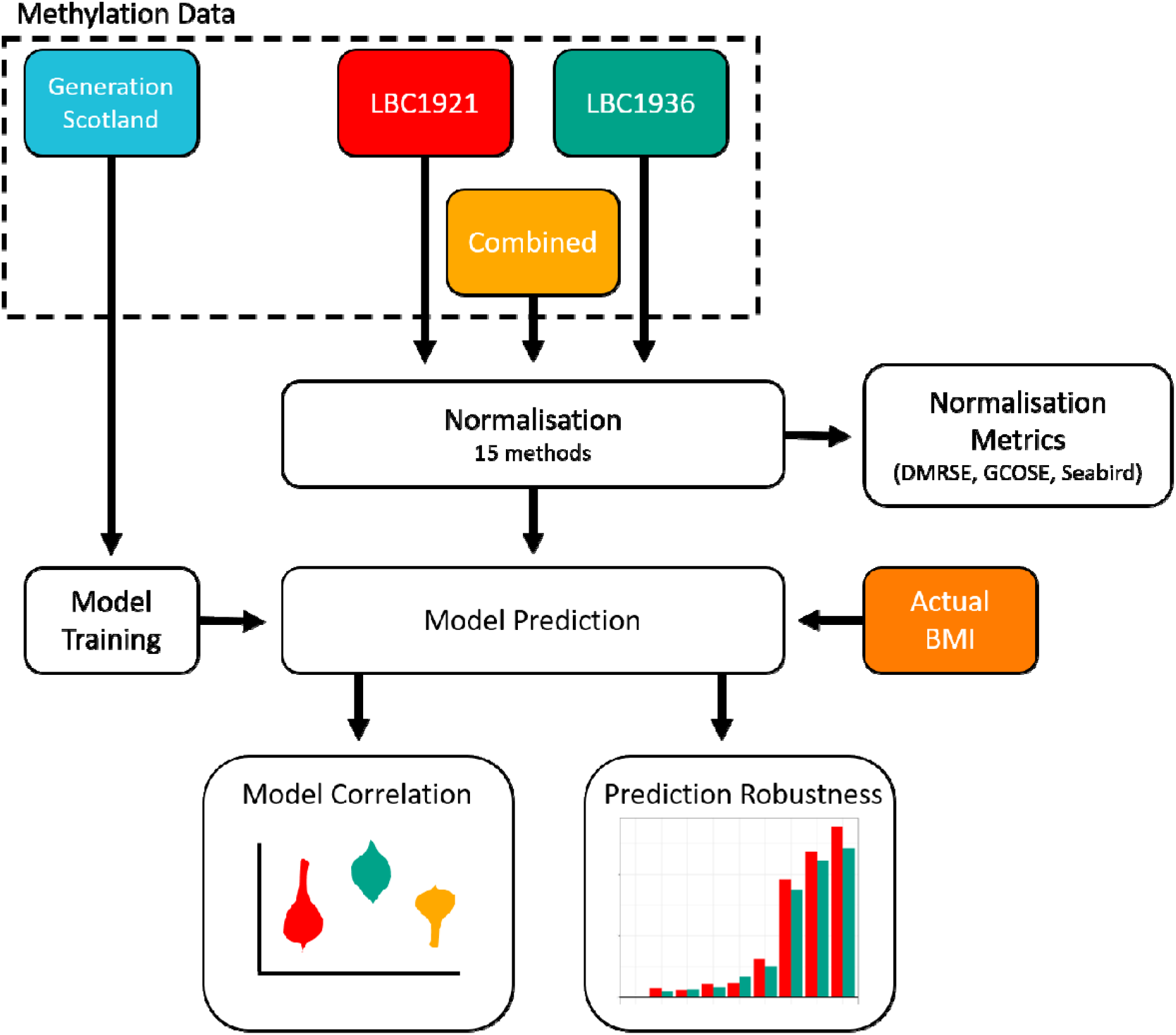
Schematic of normalisation pipeline, model training, and prediction steps.

While no single method consistently ranked highest under the three metrics considered (DMRSE, GCOSE, seabird – **Methods**), daten2 and naten performed well overall (**Supplementary Figures 1-2**). Poorer performances were observed for the Tost method, PBC and unnormalised (raw) data, based on the three wateRmelon metrics.

### DNAm-based BMI Prediction by normalisation method

Pearson correlations between observed BMI and a BMI epigenetic score (EpiScore; trained in an independent cohort of 18,413 individuals using elastic net regression – **Methods**) are shown in **Figure 2 and Supplementary Table 1** for all normalisation methods. The mean correlations were 0.34 (SD 0.02) for LBC1921 and 0.47 (SD 0.01) for LBC1936 – the maximum correlation was 0.49 (incremental R^2^ = 24.5% when comparing linear regression models of log(BMI) against age and sex with/out a BMI EpiScore) for the SWAN normalisation method in LBC1936.

**Figure 2:**
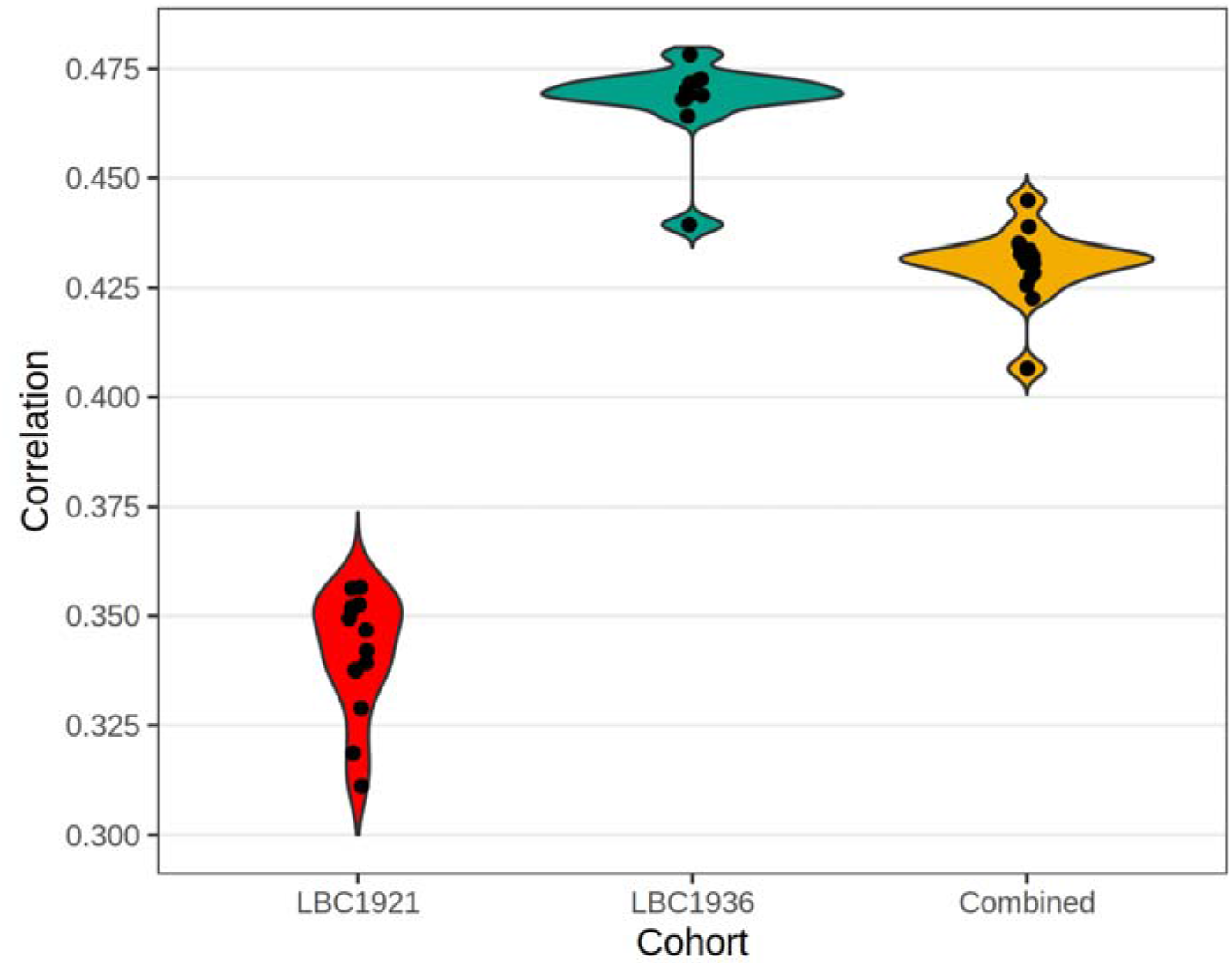
Violin plot showing the distribution of Pearson correlations between observed BMI and the BMI EpiScore across normalisation methods. Dots within the violin plots are individual normalisation method estimates.

### Prediction Robustness Metrics Assessment

While the choice of normalisation method had little effect on the EpiScore correlations with actual BMI, differences amongst the normalisation methods were present when looking into deviations in EpiScore predictions for individuals. **Figure 3** shows the mean absolute difference in EpiScores for each normalisation method. For all EpiScore predictions, the within-array methods (SWAN, Noob, PBC and BMIQ) had no mean difference because they are entirely sample-wise methods (i.e., normalisation is specific to each sample). Of the other methods, nanet was the best performing with a mean absolute difference in the predicted BMI EpiScores (0.002 units of log(BMI) adjusted for age, sex, and genetic PCs – approximately 0.01 kg/m2 after de-scaling, **Appendix 1**) for LBC1921 participants when normalised separately and together with LBC1936. **Figure 4** highlights the similarity in EpiScores for the nanet-normalised data compared to nasen (the between-array methods with the smallest and largest mean absolute differences, respectively) for LBC1936.

**Figure 3:**
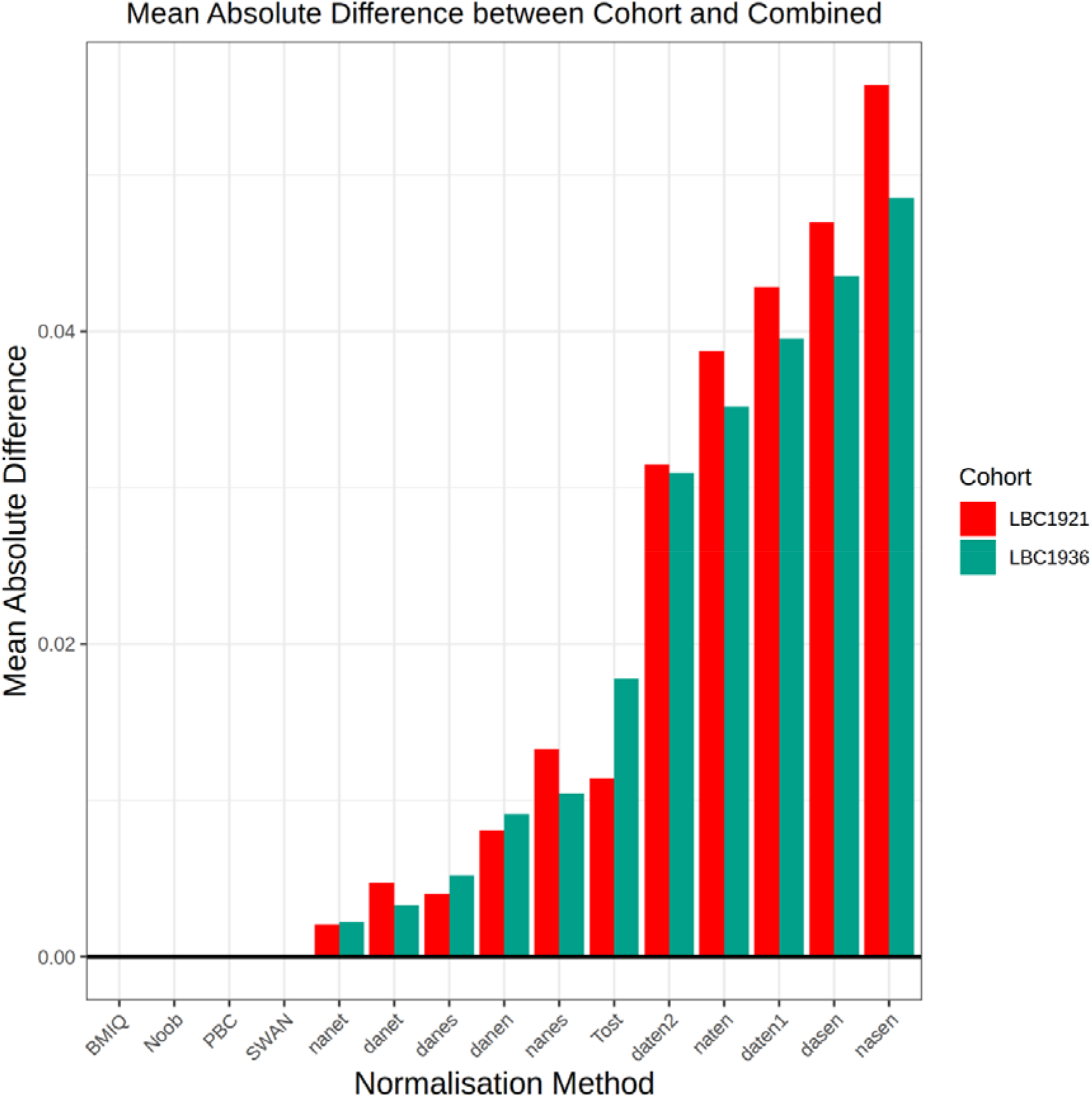
Bar plot comparing mean absolute value difference in BMI EpiScores between separately and jointly normalised cohorts.

**Figure 4:**
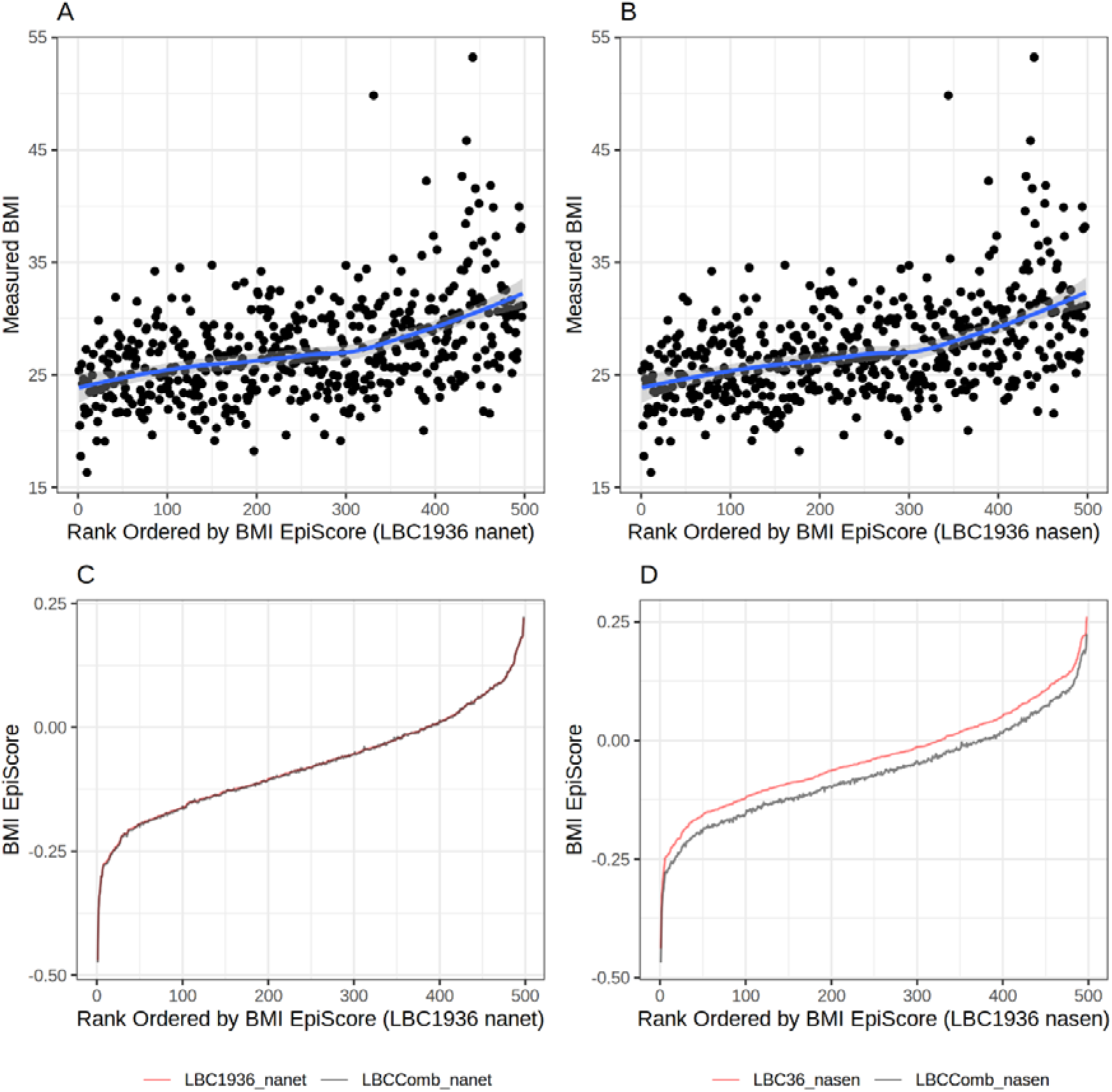
Measured BMI and BMI EpiScores in LBC1936 for the nanet (lowest MAD) and nasen (highest MAD) normalisation methods after normalising the LBC1936 data on its own and then jointly with LBC1921. Actual BMI (kg/m^2^) is plotted for individuals in ascending order of their LBC1936 EpiScore values (Panels A-B). BMI EpiScores are plotted against individuals in ascending order of their EpiScore values for the LBC1936-only dataset (red line) and together with LBC1921 (black line; Panels C-D).

## Discussion

The removal of technical noise from DNAm datasets is essential for more reliable predictions of a new individual onto a reference dataset [13]. Optimal normalisation methods should give similar predictions irrespective of whether a dataset has been normalised independently or jointly with a potential reference dataset.

Here, we showed that, for the generation of BMI EpiScores, between-array methods such as nanet and danet yield similar predictions irrespective of whether the input data was normalised with the reference dataset. As expected, within-array methods showed no differences in predictions between datasets. A range of correlations was observed between phenotypic BMI and the BMI EpiScores using these methods, with SWAN performing well in both cohorts. While within-array methods result in a zero mean absolute difference, they also guarantee that no batch-level effects are taken into account. Perhaps reflecting this, Noob yielded the second-lowest correlation between BMI and the EpiScore in LBC1921 and the jointly normalised cohorts. In contrast, nanet and danet gave both high correlations and low mean absolute differences. A common feature between these methods is the application of dye-bias correction to Type I and Type II probes together.

When projecting EpiScores for a new individual/cohort, between-array normalisation methods that don’t perform dye-bias correction (e.g., nasen, dasen, naten) gave greater differences in EpiScores when the new cohort was normalised on its own versus jointly with the second, reference population. Whether Type I or Type II probes were normalised separately or together did not seem to have a clear effect on EpiScore robustness. Technical noise due to dye bias was therefore the main cause of discrepancies for the BMI EpiScores robustness.

While no single best normalisation method is clearly highlighted, we identified strengths of normalisation techniques that will generalize well to a new dataset. Methods that do not correct for dye bias appear to have limited utility. This finding is important for the potential deployment of DNAm prediction tools in the healthcare community. Ideally, studies would not have to re-normalise a dataset every time new volunteers or patients are entered, which is both time consuming and computationally expensive.

There are also some general limitations around prediction that need to be considered. Firstly, the BMI EpiScore was trained and tested in Scottish populations. Its correlation with observed BMI differed substantially across the two LBC studies, despite minimal heterogeneity in age and background. Differences are also likely to exist when EpiScores are applied to more diverse populations (e.g., wider age ranges, different social backgrounds and ancestries), though the tightly-matched age range across both test cohorts reported here is valuable in that cross-cohort comparisons were not confounded by age differences. EpiScore predictors for more complex traits or diseases risk scores could also be trained [2].

Most existing EpiScore analyses have focused on relative differences within a cohort or analysis batch [1]. By applying and comparing a variety of normalisation approaches, we suggest individuals or cohorts could be reliably projected onto a reference panel. This will enable users to generate methylation-based scores and risk percentiles for a variety of traits and diseases.

## Methods

### Lothian Birth Cohorts (LBC) of 1921 and 1936 -DNAm Quality Control

DNAm data was assessed using the Illumina 450k array for 499 individuals from the LBC1936 and 436 individuals from the LBC1921 [12]. Prior to normalisation, each sample went through a number of filtering checks. P-values to quantify signal reliability (detection P-values) were computed for each CpG probe. Probes which had more than 1% of samples with a P-value greater than 0.05 were removed (7,366 LBC1921 probes were removed, 1,495 LBC1936 probes were removed) Individual samples where more than 1% of probes had a detection P-value greater than 0.05 were removed (49 LBC1921 samples, 1 LBC1936 sample removed) Finally, we removed probes with a bead count of less than three in more than 5% of samples (191 LBC1921 probes removed, 362 LBC1936 probes removed). Following quality control, there were 445,962 remaining probes common across all datasets. There were 387 individuals remaining in the LBC1921 cohort and 498 individuals remaining in the LBC1936 cohort. The combined cohort had 885 individuals.

### Normalisation Methods

Fifteen normalisation methods from the Minfi and WateRmelon packages [7,10,11] were applied to LBC1921, LBC1936, and the combined LBC dataset with the pipeline depicted in **Figure 1**.

WateRmelon is an R package that implements several QN methods with systematic nomenclature described in [11]. Methods which start with a ‘d’ apply background adjustment (‘n’ indicates no adjustment). The third letter specifies whether between-array normalisation was performed to Type I and II probes separately (‘s’), together (‘t’), or not at all (‘n’). The final letter indicates whether dye-bias correction was applied to Type I and II probes separately (‘s’), together (‘t’), or not at all (‘n’). A description of the difference between normalisation methods is shown in **Supplementary Table 2**.

Minfi is an R package [14] that contains two additional normalisation techniques, Noob and SWAN. Normal-exponential out-of-bound (Noob) is a within-sample background correction method with dye-bias normalisation for DNAm arrays [10]. Noob uses a normal-exponential convolution method to estimate background distributions by measuring non-specific fluourescence based on out-of-band Type I (i.e. probes in the opposite colour channel - Cy3 vs Cy5), Subset-quantile within array normalisation (SWAN) consists of two steps [7]. The first step takes a subset of probes, defined to be biologically similar based on CpG content, and determines an average quantile distribution from this subset. The second step adjusts the intensities of the remaining probes by linear interpolation onto the distribution of the subset probes.

In addition to the wateRmelon and Minfi functions, we applied 3 widely-cited methods in our comparison: BMIQ, peak-based correction and subset quantile normalisation [8, 9, 15]. BMIQ (a within-array method) first fits a 3-state beta mixture model (0%. 50% and 100% methylation) for Type I and Type II probes separately, in which probes are assigned to the state with maximum probability. This is followed by normalisation of Type II probes to the distributiuons of Type I probes in the same group. Peak-based correction independently estimates M-value peaks for Type I and Type II probes, followed by rescaling of the Type II assays to match the estimates obtained for Type I assays. Subset quantile normalisation (Tost method), normalises signal from Type II assays based on a set of Type I “anchor” probes, which are considered to be more reliable and stable.

### Normalisation assessment metrics

Three previously published performance metrics were considered [11]. Differentially Methylated Regions Standard Error (DMRSE) measures the variation at sites defined as uniparentally methylated regions with an expected β value of 0.5. The standard error is computed by dividing the standard deviation of differentially methylated region β values by the square root of the number of samples. Genotype Combined Standard Error (GCOSE) examines highly polymorphic SNPs which have three genotypes: heterozygous or homozygous with the major or minor allele. This metric clusters observations into the three groups based on genotype and computes a mean-squared error for each cluster, then averages the three means. Finally, the Seabird metric computes the area under the curve (AUC) for a predictor trained on sex differences on the X chromosome, of which one is hypermethylated in females. Each of the normalisation metrics was ranked on each of the three metrics; the ranks were then averaged to compute a mean overall rank.

### DNAm predictor of BMI

A DNAm predictor of body mass index (BMI) was derived using elastic net penalised regression (α = 0.5) on 18,413 participants from the Generation Scotland study [16]. The lambda value that minimised the mean error in a 10-fold cross validation analysis resulted in a weighted linear predictor containing 3,506 CpGs (see **Supplementary Table 3**). As the Generation Scotland DNAm resource was generated using the EPIC array, CpGs were first subset to the 445,962 sites that were common to the 450k array and that passed QC in the LBC analyses. They were further pruned to the 200,000 most variable CpG features (ranked by standard deviation) to avoid a memory allocation error in the elastic net model. R’s biglasso package was used to implement an elastic net regression model [17–19]. The input to the model was a 200,000 × 18,413 matrix containing the CpG M-values for each individual. The target variable was the residuals from a linear regression model of log(BMI) adjusted for age, sex and 10 genetic principal components. The distribution of BMI in the two LBC studies and Generation Scotland are presented in **Supplementary Figure 3**.

### Prediction and Robustness

Predictions of BMI were performed on both LBC datasets and the combined LBC dataset. An individual’s BMI was predicted by weighting their CpG values by the CpG weights from the Generation Scotland elastic net model. Overall model prediction performance was evaluated by Pearson’s correlation coefficient.

Prediction robustness measures a normalisation method’s invariance to datasets being normalised independently, or jointly with another dataset. Robustness was calculated as the mean absolute difference between the independent and joint predictions across all individuals. The goal is to identify how the test datasets behave when predictions are made using data normalised jointly or separately. Small mean differences indicate normalisation methods that provide similar outputs irrespective of the data being normalised separately or together. Normalisation methods with large mean absolute differences result in inconsistent predictions depending on whether new individuals are normalised jointly with previous data or not.

## Availability of Data

According to the terms of consent for Generation Scotland (GS) participants, access to data must be reviewed by the GS Access Committee. Applications should be made to.

Lothian Birth Cohort data are available on request from the Lothian Birth Cohort Study, University of Edinburgh (https://www.ed.ac.uk/lothian-birth-cohorts/dataaccess-collaboration). Lothian Birth Cohort data are not publicly available due to them containing information that could compromise participant consent and confidentiality.

All code is available with open access at the following GitHub repository: https://github.com/marioni-group/DNAm_EpiScore_Projections

## Data Availability

Lothian Birth Cohort data are available on request from the Lothian Birth Cohort Study, University of Edinburgh (https://www.ed.ac.uk/lothian-birth-cohorts/data-access-collaboration). Lothian Birth Cohort data are not publicly available due to them containing information that could compromise participant consent and confidentiality.

## Acknowledgements

This research was funded in whole, or in part, by the Wellcome Trust (104036/Z/14/Z and 216767/Z/19/Z). For the purpose of open access, the author has applied a CC BY public copyright licence to any Author Accepted Manuscript version arising from this submission.

Generation Scotland received core support from the Chief Scientist Office of the Scottish Government Health Directorates [CZD/16/6] and the Scottish Funding Council [HR03006] and is currently supported by the Wellcome Trust [216767/Z/19/Z]. Genotyping of the GS samples was carried out by the Genetics Core Laboratory at the Edinburgh Clinical Research Facility, University of Edinburgh, Scotland and was funded by the Medical Research Council UK and the Wellcome Trust (Wellcome Trust Strategic Award “STratifying Resilience and Depression Longitudinally” (STRADL) Reference 104036/Z/14/Z).

The authors thank all LBC study participants and research team members who have contributed, and continue to contribute, to the ongoing LBC study. The LBC1936 is supported by the Biotechnology and Biological Sciences Research Council, and the Economic and Social Research Council [BB/W008793/1], Age UK (Disconnected Mind project), the Milton Damerel Trust, and the University of Edinburgh. The LBC1921 was supported by the Biotechnology and Biological Sciences Research Council [SR176], the Chief Scientist Office of the Scottish Government [CZB/4/505; ETM/55], the Royal Society and the Medical Research Council [R42550]. Methylation typing was supported by the Centre for Cognitive Ageing and Cognitive Epidemiology (Pilot Fund award), Age UK, The Wellcome Trust Institutional Strategic Support Fund, The University of Edinburgh, and The University of Queensland. CM, TG, and BR are supported by the United Kingdom Research and Innovation (grant EP/S02431X/1), UKRI Centre for Doctoral Training in Biomedical AI at the University of Edinburgh, School of Informatics. REM is supported by Alzheimer’s Society major project grant AS-PG-19b-010.

## Conflicts of interest

REM is a scientific advisor to the Epigenetic Clock Development Foundation and Optima Partners. RFH is a scientific advisor to Optima Partners. LM has received payment from Illumina for presentations and consultancy.

---

**Supplementary Table 1:**
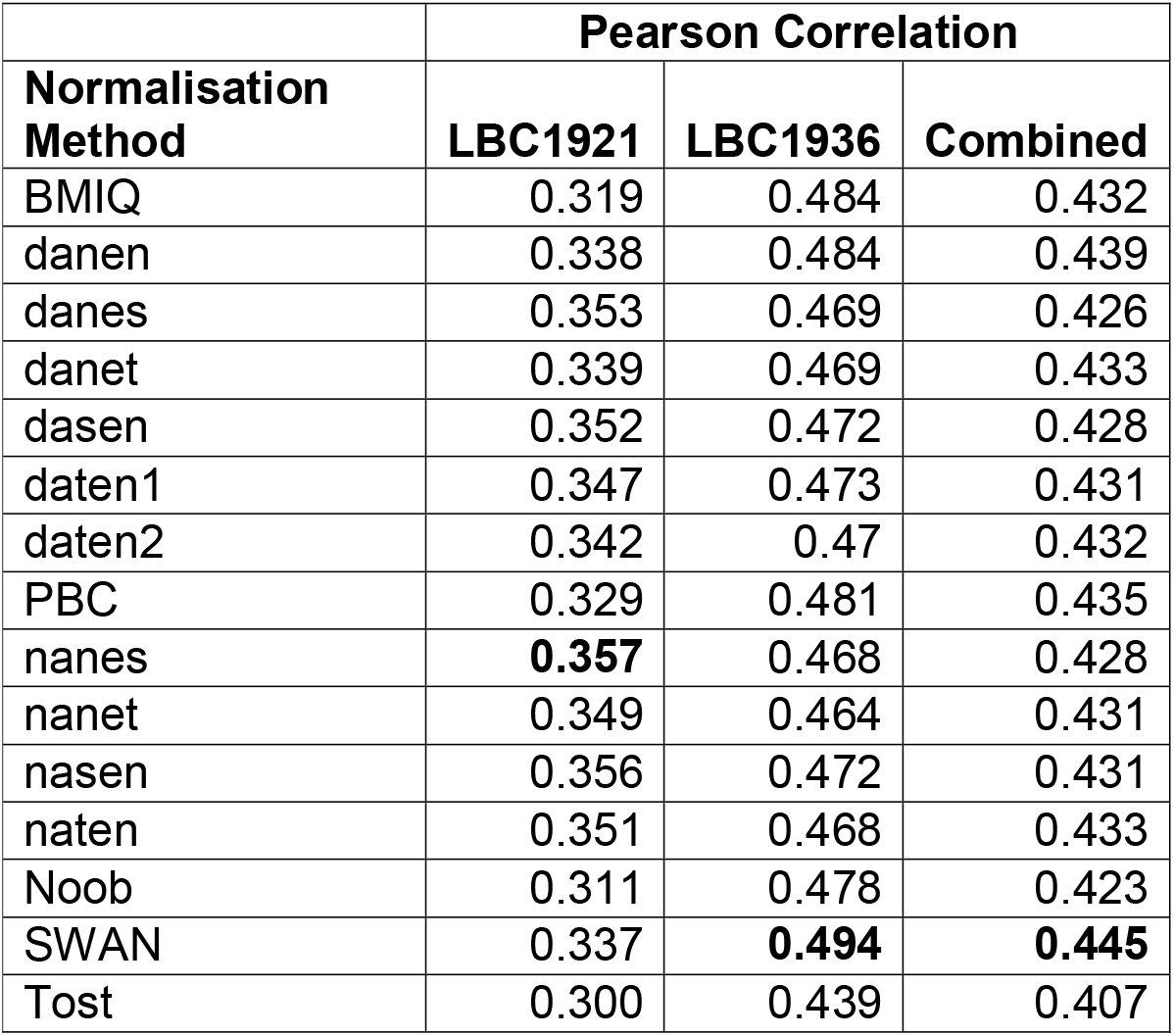
Correlations between measured BMI and the BMI EpiScore in LBC1921, LBC1936 and LBC1921 + LBC1936 (Combined) for every normalisation method. The largest value in each column is highlighted in bold.

**Supplementary Table 2.** WateRmelon normalisation method descriptions.

| Description of Normalisation Methods |  |  |  |
| --- | --- | --- | --- |
| Normalisation | Background Adjustment | Between-Array normalisation | Dye-Bias Correction |
| naten | None | Type I/II together | None |
| nanet | None | None | Type I/II together |
| nanes | None | None | Type I/II separately |
| nasen | None | Type I/II separately | None |
| danet | Adjusted | None | Type I/II together |
| danes | Adjusted | None | Type I/II separately |
| dasen | Adjusted | Type I/II separately | None |

**Supplementary Table 3:** 2023-03-03_Table_S3.csv

**Supplementary Figure 1:**
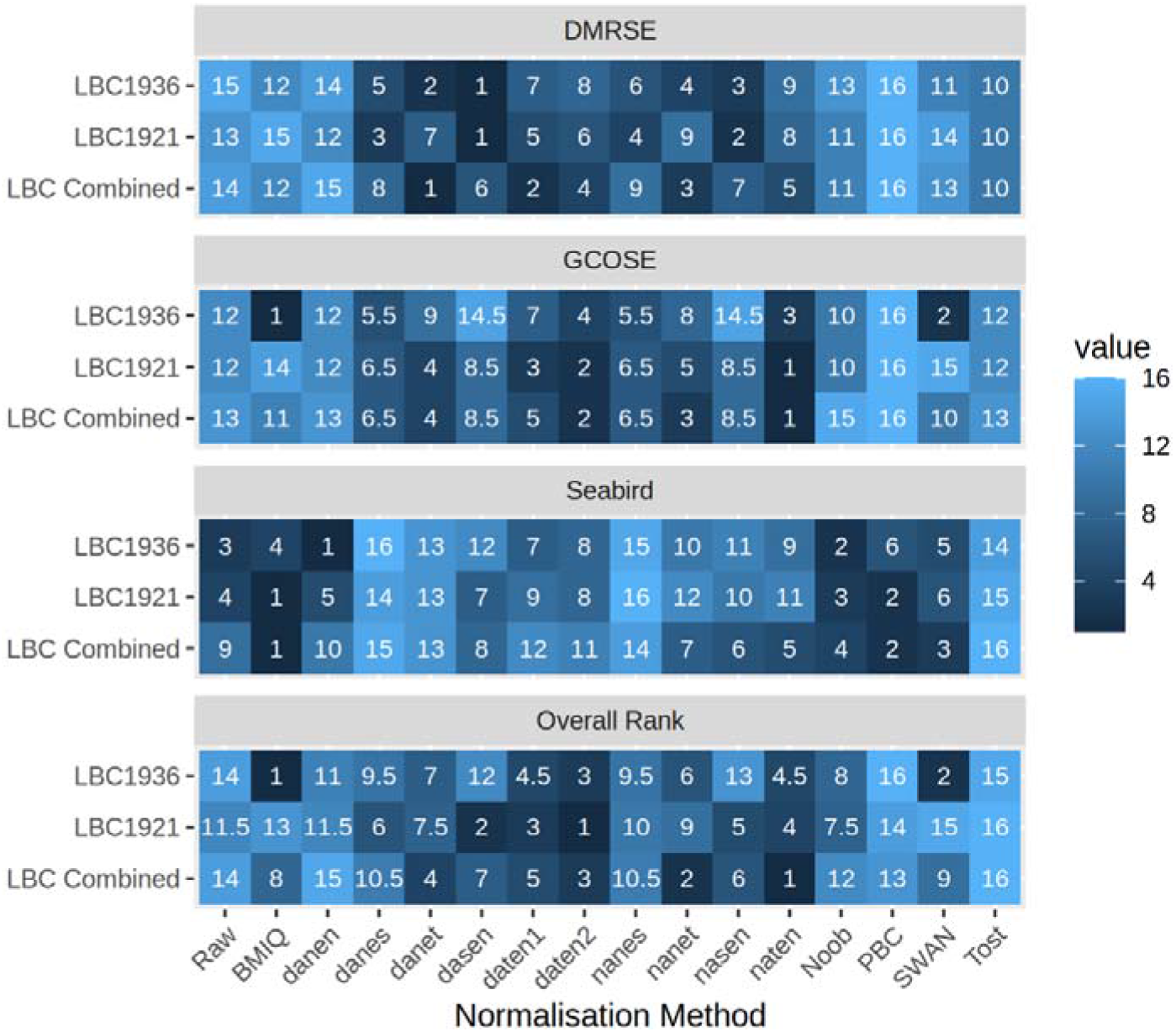
Heatmaps of normalisation method ranks for the DMRSE, GCOSE and Seabird metrics.

**Supplementary Figure 2:**
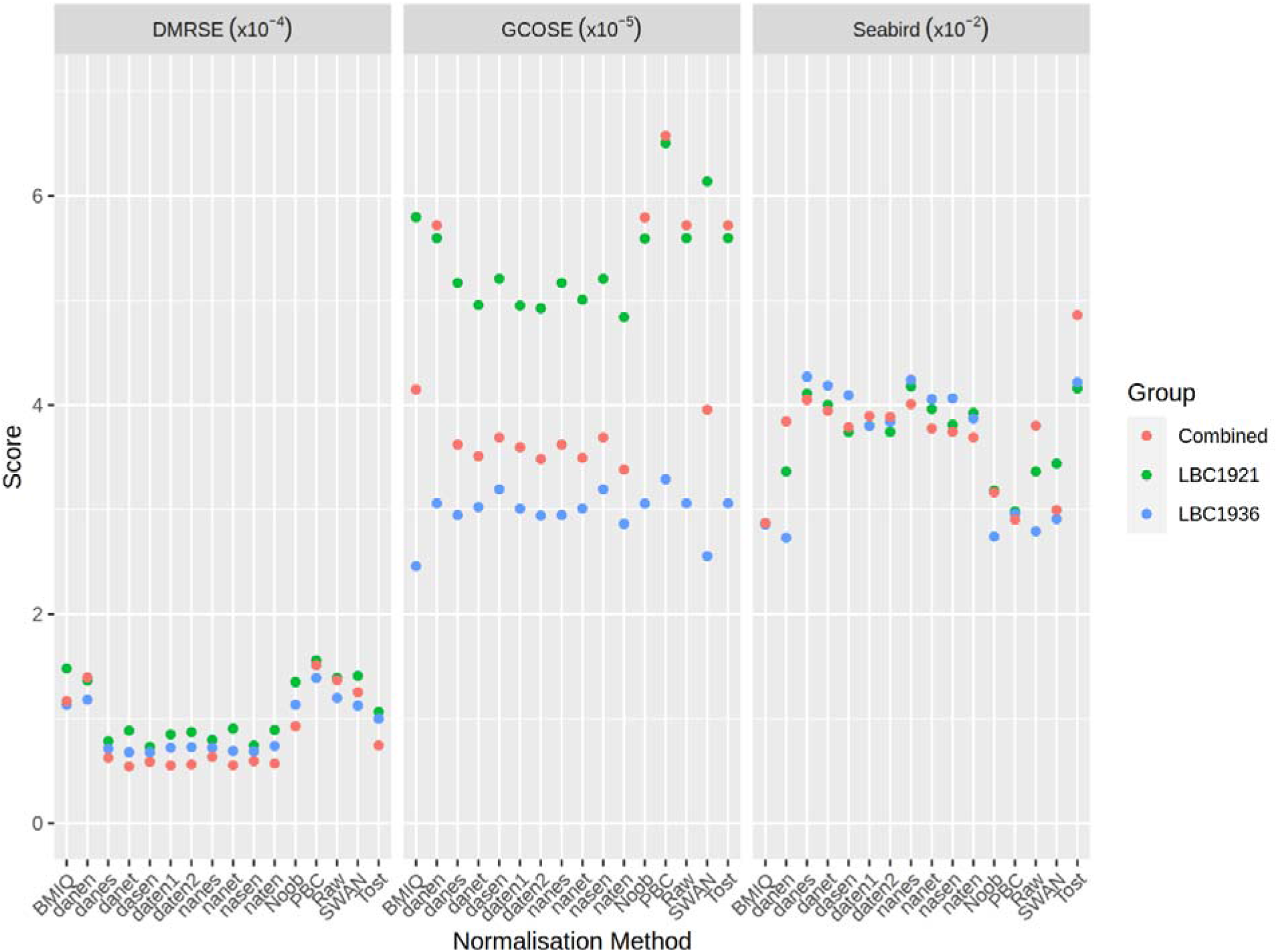
Individual scores for the DMRSE, GCOSE and Seabird metrics across normalisation methods applied to LBC1921, LBC1936 and both cohorts combined.

**Supplementary Figure 3:**
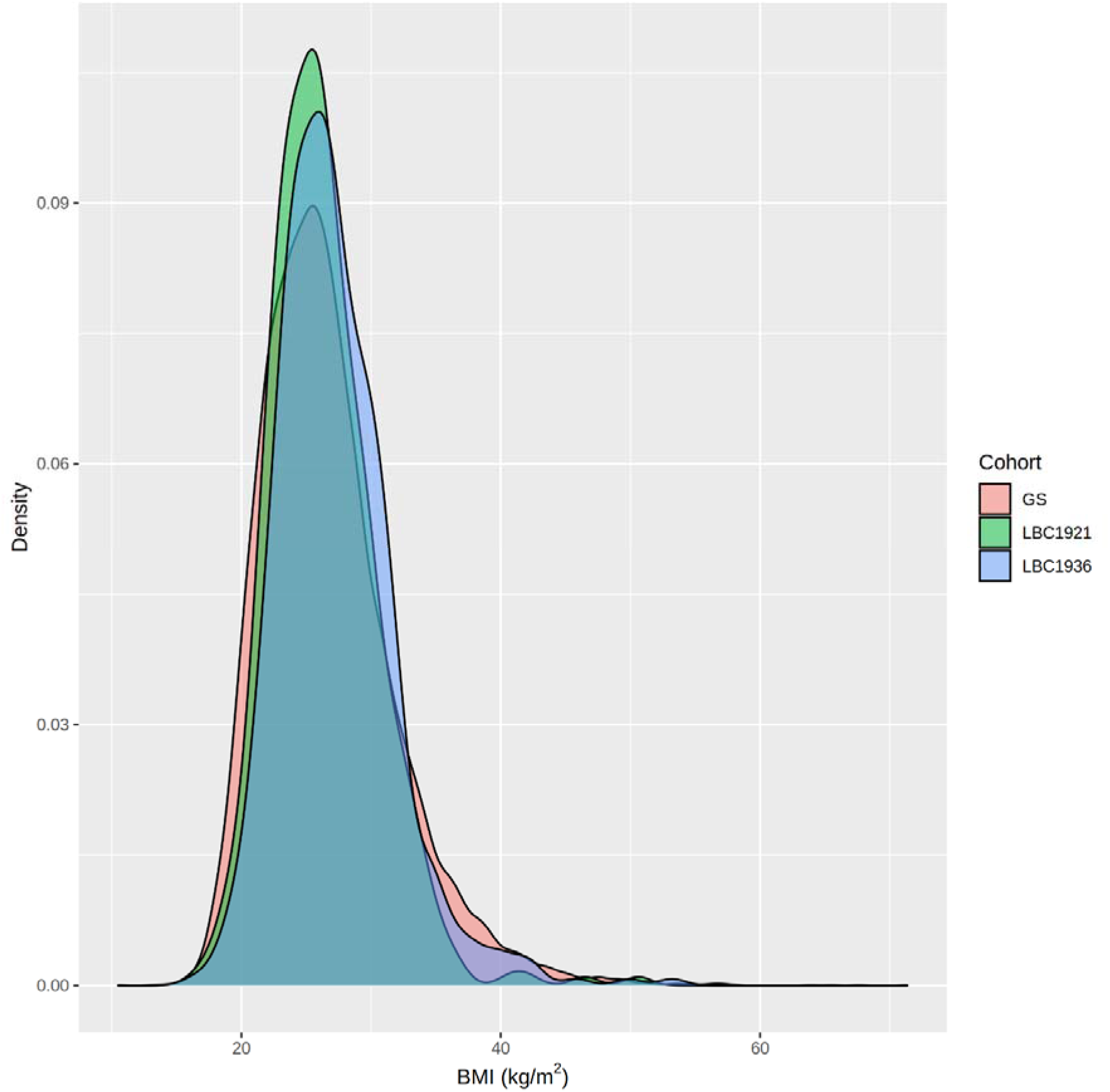
Density plot of BMI (kg/m^2^) in the Lothian Birth Cohort 1921, the Lothian Birth Cohort 1936, and Generation Scotland.

## Appendix 1: Converting BMI EpiScores back to the original (BMI kg/m^2^) scale

The following formula was used to rescale the BMI EpiScores:

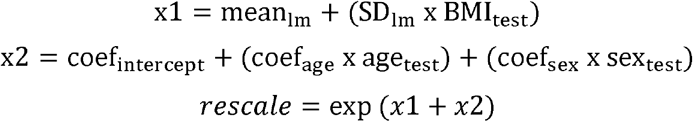

Where *lm* is the residual from the model log(BMI) ∼ age + sex in the training set (Generation Scotland), *test* is the corresponding test sample (LBC1921, LBC1936 or Combined), and *coef* is the set of coefficients from the model *lm*.

## Notes

### Author Declarations

All components of GS received ethical approval from the NHS Tayside Committee on Medical Research Ethics (REC Reference Number: 05/S1401/89). GS has also been granted Research Tissue Bank status by the East of Scotland Research Ethics Service (REC Reference Number: 20-ES-0021), providing generic ethical approval for a wide range of uses within medical research. Ethical approval for the LBC1921 and LBC1936 studies was obtained from the Multi-Centre Research Ethics Committee for Scotland (MREC/01/0/56) and the Lothian Research Ethics committee (LREC/1998/4/183; LREC/2003/2/29). In both studies, all participants provided written informed consent. These studies were performed in accordance with the Helsinki declaration.

